# Music listening for chronic pain management: a systematic review, meta-analysis, and evaluation of intervention reporting quality

**DOI:** 10.64898/2026.07.08.26357000

**Authors:** Jèssica Garrido-Pedrosa, Mᵃ Teresa Sáez Vasco, Lorena Zapata, Maria F. Porto Torres, Rocío Valenzuela, Antoni Rodríguez-Fornells, Víctor Fernández-Dueñas, Jennifer Grau-Sánchez

## Abstract

**Background:** Chronic pain is a multidimensional condition that often persists despite conventional treatment and adversely affects multiple domains of daily life. Music listening has emerged as a promising non-pharmacological intervention, with accumulating evidence supporting its beneficial effects on pain and associated psychological outcomes. However, despite growing evidence of efficacy, the translation of music listening into routine clinical practice remains limited, partly because intervention reporting has received comparatively little attention.

**Objective:** To evaluate the effectiveness of music listening interventions for chronic pain and systematically assess the methodological quality and completeness of intervention reporting to identify barriers to reproducibility and clinical implementation.

**Methods:** Systematic searches were conducted in PubMed, Cochrane Library, CINAHL, and Web of Science through June 2025, with no date restrictions on publication. Randomized controlled trials involving adults with chronic pain receiving music listening interventions were included. Two independent reviewers screened studies, extracted data, and assessed risk of bias. Intervention reporting was evaluated using the TIDieR checklist, and a random-effects meta-analysis was performed for pain intensity outcomes.

**Results:** Ten RCTs involving 538 participants were included. Music listening interventions varied substantially in delivery, duration, and music selection procedures, reflecting considerable heterogeneity in intervention design. Most studies reported significant improvements in pain and psychological outcomes. Meta-analysis of eight trials (10 effect estimates), demonstrated a moderate reduction in pain intensity (SMD = −0.53, 95% CI: −0.96 to −0.11, p = 0.014; I² = 76.2%). Although intervention rationale and procedures were generally well described, reporting of intervention modifications, treatment fidelity, and adherence was frequently incomplete. These reporting deficiencies may compromise reproducibility and limit translation into clinical practice.

**Conclusions:** Music listening appears to be a safe, accessible, and scalable non-pharmacological intervention for chronic pain management, with benefits extending beyond pain reduction to psychological wellbeing, quality of life, and functioning. However, incomplete reporting of key intervention components may limit reproducibility and hinder clinical implementation. Future trials should adopt standardized and transparent reporting standards to facilitate implementation into clinical practice.

## 1. Introduction

Chronic pain, affecting an estimated 10% to 40% of the global adult population, is one of the most prevalent and debilitating conditions worldwide (Cohen et al., 2021). It is more common among women, older adults, and socially vulnerable populations, and is one of the leading causes of disability and healthcare use (Breivik et al., 2013; Fishman, 2007). Beyond the personal burden experienced by patients, chronic pain is associated with an increased risk of depression, Alzheimer’s disease, substance abuse, and suicide (Balit et al., 2024; Mullins et al., 2023; Racine, 2018; Vadivelu et al., 2018; Wang et al., 2024), and its high treatment costs and prolonged medication use contribute substantially to healthcare expenditures (Guy et al., 2025; Stubhaug et al., 2024).

Defined as persistent or recurring pain lasting more than three months, chronic pain is a multidimensional condition encompassing biological, psychological and social factors (Bonazza et al., 2024; Howlin & Rooney, 2020; Metzner et al., 2022). Accordingly, the International Classification of Diseases (ICD-11) classifies chronic pain into distinct syndromes based on their underlying etiology, including chronic cancer pain, neuropathic pain, musculoskeletal pain, post-traumatic or postsurgical pain, visceral pain, headache and orofacial pain, and primary pain. Regardless of its underlying cause, chronic pain is influenced by dynamic reciprocal interactions between biological, psychological, and social factors (Edwards et al., 2016; Gatchel et al., 2007), and its impact extends well beyond physical discomfort, disrupting emotional, social, and occupational functioning and frequently co-occurring with anxiety and depression (Hadi et al., 2019; Lerman et al., 2015).

Pharmacological treatments, guided by the World Health Organization’s analgesic ladder, remain the primary approach for managing chronic pain. However, reliance on medications, particularly opioids, carries risk of adverse effects and addiction (Chou et al., 2017; Yang et al., 2020). The opioid crisis has underscored the urgent need for non-pharmacological interventions that address the multidimensional nature of chronic pain while reducing the risks associated with prolonged medication use (Saloner et al., 2018; Vadivelu et al., 2018). Among these, music-based therapies represent a non-invasive, safe, cost-effective, and accessible strategy that has gained increasing attention (Garza-Villarreal et al., 2017). Music listening, in particular, can be easily integrated into daily life and shows promising therapeutic potential across diverse settings and populations (Sihvonen et al., 2022).

The analgesic potential of music is supported by plausible neurobiological mechanisms. Music modulates brain networks involved in the affective and cognitive processing of pain and engages the mesolimbic reward system through dopaminergic signalling (Ferreri et al., 2019; Pando-Naude et al., 2019; Salimpoor et al., 2011). These mechanisms may be particularly relevant in chronic pain, where dysfunction of reward-related circuits has been linked to anhedonia, depression, and reduced motivation (Serafini et al., 2020). These neurobiological mechanisms likely interact with psychological processes such as distraction, relaxation, and emotional regulation. Together, these interacting mechanisms provide a plausible biological rationale for the therapeutic effects of music listening.

Reflecting this interest, a growing body of evidence has accumulated. Several systematic reviews and meta-analyses have reported beneficial effects of music interventions on pain and emotional outcomes (Cepeda et al., 2006; Garza-Villarreal et al., 2017; Hsu et al., 2022; Lee, 2016; Martin-Saavedra et al., 2018), including a recent meta-analysis confirming significant reductions in pain and depression in chronic pain populations (Chen et al., 2025). A recent scoping review of 63 studies similarly concluded that music offers benefits not only for pain but also for emotional regulation, anxiety, depression, and social dimensions of the chronic pain experience (Cournoyer Lemaire & Perreault, 2024). The available evidence consistently supports the effectiveness of music listening for reducing pain and improving psychological wellbeing. However, this accumulating evidence has not translated into clear clinical recommendations or standardized protocols. Reported effect sizes vary widely, heterogeneity across trials is typically high, and there is little consensus on how these interventions should be delivered in practice. This persistent gap between growing evidence and clinical implementation suggests that further efficacy trials alone are unlikely to advance the field.

The central challenge has shifted from demonstrating efficacy to identifying how music listening interventions can be standardized, replicated, and implemented across clinical settings. Music listening constitutes a complex intervention rather than a single, uniform treatment. Its delivery depends on multiple components: the procedure by which music is selected, the dose and schedule of listening, the mode and setting of delivery, the degree of participant involvement, and the monitoring of adherence and treatment fidelity. When these components are described incompletely, it becomes difficult to determine which elements contribute to benefit, to compare findings across trials, or to replicate and implement successful protocols in practice. Transparent and complete intervention reporting is therefore not merely an editorial requirement but a prerequisite for translating efficacy into clinical practice.

Standardized frameworks have been developed precisely to address this problem. The Template for Intervention Description and Replication (TIDieR) checklist (Hoffmann et al., 2014) provides a structured basis for describing the components of an intervention in sufficient detail to permit replication. Although widely adopted across health research, such frameworks have rarely been used to evaluate the reporting quality of music listening interventions in chronic pain. Existing reviews have focused almost exclusively on estimating treatment effects and have devoted comparatively little attention to how interventions are described and reported. As a result, the field lacks a systematic account of whether the available trials report enough to be reproduced and implemented, and of which components are most often missing. Beyond music listening itself, these limitations reflect a broader challenge in evaluating complex non-pharmacological interventions, where active components are often difficult to define and report consistently (Grau-Sánchez et al., 2022a; Rodríguez-Fornells et al., 2025a; Skov & Nadal, 2025).

Improving the reporting of music listening interventions may therefore represent one of the most direct strategies for accelerating their translation into routine clinical care. Accordingly, this systematic review and meta-analysis pursues two complementary aims. First, we synthesize the evidence on the effectiveness of music listening for chronic pain, examining its effects on pain intensity and on anxiety, depression, mood, quality of life, and functionality, and quantifying the pooled effect on pain through meta-analysis. Second, we systematically assess the methodological quality and the completeness of intervention reporting of the available trials, using the TIDieR checklist together with risk-of-bias appraisal. By integrating evidence synthesis with a structured evaluation of reporting quality, this review seeks not only to synthesize the available evidence but also to identify the methodological barriers limiting reproducibility and implementation.

## 2. Methods

We conducted a systematic review and meta-analysis to evaluate the effectiveness of music listening interventions on chronic pain management. The review protocol was prospectively registered in the International Prospective Register of Systematic Reviews (PROSPERO; registration number CRD42024493588), and the review was conducted and reported in accordance with the Preferred Reporting Items for Systematic Reviews and Meta-Analyses (PRISMA) 2020 statement (Page et al., 2021). Detailed search strategies, eligibility criteria, and the data extraction sheet are provided in the Supplementary Material (**Tables S1-S3**).

### 2.1. Information sources and search strategy

A systematic literature search was performed in PubMed (via MEDLINE), the Cochrane Central Register of Controlled Trials (CENTRAL; Cochrane Library), the Cumulative Index to Nursing and Allied Health Literature (CINAHL; EBSCOhost), and Web of Science (Clarivate). Search strategies combined controlled vocabulary and free-text terms related to chronic pain, music listening interventions, and relevant clinical outcomes, including pain, anxiety, depression, mood, quality of life, and functionality. Searches were limited to articles published in English or Spanish. To maximize completeness, the reference lists of all eligible studies were screened manually for additional relevant publications. No restrictions were applied regarding publication date, and no additional sources, such as trial registries or conference proceedings, were searched. The initial search was conducted in September 2024 and updated through June 2025. The complete search strategies for each database are available in the registered PROSPERO protocol (CRD42024493588). The full electronic search strategy for each database is provided in **Supplementary Table S1**.

### 2.2. Eligibility criteria

Studies were eligible if they: (1) were randomized controlled trials (RCTs); (2) included adults (≥18 years) with any type of chronic pain, defined as persistent or recurrent pain lasting longer than three months; (3) evaluated the effectiveness of music listening interventions; and (4) reported at least one of the following outcomes: pain intensity, anxiety, depression, mood, quality of life, or functionality in activities of daily living. Only articles published in English or Spanish were considered. Music listening interventions were defined as receptive interventions in which participants attentively listened to music, including protocols combined with relaxation or guided imagery. Interventions involving active music-making (e.g., singing, playing instruments, or moving to music) were excluded to isolate the effects of receptive music listening. No restrictions were applied regarding the type of comparator, clinical setting, or publication date. The complete eligibility criteria, structured according to the PICOS framework, are summarized in **Supplementary Table S2**.

### 2.3. Selection process

Two reviewers (JGP and MTSV) independently screened all records identified through the literature search according to the predefined eligibility criteria. Titles and abstracts were initially assessed, and full-text articles were retrieved whenever eligibility could not be determined from the available information or when the study appeared potentially relevant. Duplicate records were identified and removed using Mendeley reference management software, followed by manual verification.

The full texts of all potentially eligible studies were then independently assessed by both reviewers to determine final inclusion in the systematic review. Any disagreements at any stage of the selection process were resolved through discussion, and when consensus could not be reached, a third reviewer (JGS) acted as an adjudicator.

### 2.4. Data collection process

Data were independently extracted by two reviewers (JGP and MTSV) using a standardized data extraction form. The following information was collected from each included study: study characteristics (authors, year of publication, country, journal, and study design); participant characteristics (sample size, group allocation, age, sex, chronic pain condition, and eligibility criteria); intervention characteristics (duration, frequency, session length, music type, mode of music delivery, and comparator); and outcome measures together with the main study findings. The complete data extraction sheet, including the populated table for all included studies, is provided in **Supplementary Table S3**. Any discrepancies between reviewers were resolved through discussion or, when necessary, by consultation with a third reviewer (JGS).

When relevant information was missing, unclear, or insufficiently detailed, the corresponding authors were contacted to request additional information.

### 2.5. Quality of intervention reporting assessment

The completeness of intervention reporting was evaluated using the Template for Intervention Description and Replication (TIDieR) checklist (Hoffmann et al., 2014). This 12-item checklist provides a structured framework for assessing whether interventions are described in sufficient detail to permit replication, including their rationale, procedures, delivery, dose, tailoring, modifications, and the assessment of treatment adherence and fidelity.

Beyond evaluating individual studies, the TIDieR assessment allowed us to identify which intervention components were consistently reported across the available evidence, which were systematically underreported, and the reporting deficiencies most likely to limit reproducibility and clinical implementation.

The assessment was performed independently by two reviewers (JGP and MTSV). Any discrepancies were resolved through discussion or, when necessary, by consultation with a third reviewer (JGS).

### 2.6. Study risk of bias assessment

The methodological quality of the included studies was independently assessed by two reviewers (JGP and MTSV) using the Cochrane Risk of Bias tool (RoB 1), following the recommendations of the Cochrane Handbook for Systematic Reviews of Interventions (Higgins et al., 2024). Risk of bias assessments were performed using Review Manager 5 (RevMan 5).

The following domains were evaluated: random sequence generation, allocation concealment, blinding of participants and personnel, blinding of outcome assessment, incomplete outcome data, selective outcome reporting, and other potential sources of bias. Each domain was judged as having a low, high, or unclear risk of bias according to the Cochrane Handbook criteria. Disagreements between reviewers were resolved through discussion or, when necessary, by consultation with a third reviewer (JGS). The overall methodological quality of each study was determined from the combined assessment across all domains.

### 2.7. Statistical analysis of pain outcomes

A meta-analysis was performed to evaluate the effect of music listening interventions on pain intensity. Pain intensity data were extracted from each study, and when change scores were not reported, mean changes and their corresponding standard deviations (SDs) were derived from pre-and post-intervention values whenever possible. Standardized mean differences (SMDs) with 95% confidence intervals (CIs) were calculated to account for the use of different pain assessment scales across studies and pooled using a random-effects model, given the expected clinical and methodological heterogeneity among interventions and study populations.

For trials including more than one eligible music listening intervention arm (Raglio et al., 2023; Siedliecki & Good, 2006), each intervention arm was entered as a separate comparison against the shared control group, resulting in ten effect estimates from eight randomized controlled trials. Trials were excluded from the quantitative synthesis when pain intensity outcomes could not be incorporated into the meta-analysis, either because intensity-related outcomes were not reported (Torres et al., 2018), or because only a single post-intervention assessment was available (Lin et al., 2020).

Statistical heterogeneity was assessed using Cochran’s Q test and quantified with the I² statistic, with values of approximately 25%, 50%, and 75% representing low, moderate, and high heterogeneity, respectively. All analyses were performed using Review Manager 5 (RevMan 5; Cochrane Collaboration) and the metafor package in R.

## 3. Results

### 3.1. Study selection

The literature search identified 1,801 records, including 1,799 retrieved through database searches and two identified by manual screening of reference lists. After removing 463 duplicates, 1,338 records underwent title and abstract screening, leading to the exclusion of 1,216 articles. Full texts of the remaining 122 studies were assessed for eligibility, of which 10 randomized controlled trials met the inclusion criteria and were included in the systematic review. The study selection process and reasons for exclusion are presented in the PRISMA flow diagram (**Figure 1**).

**Figure 1.**
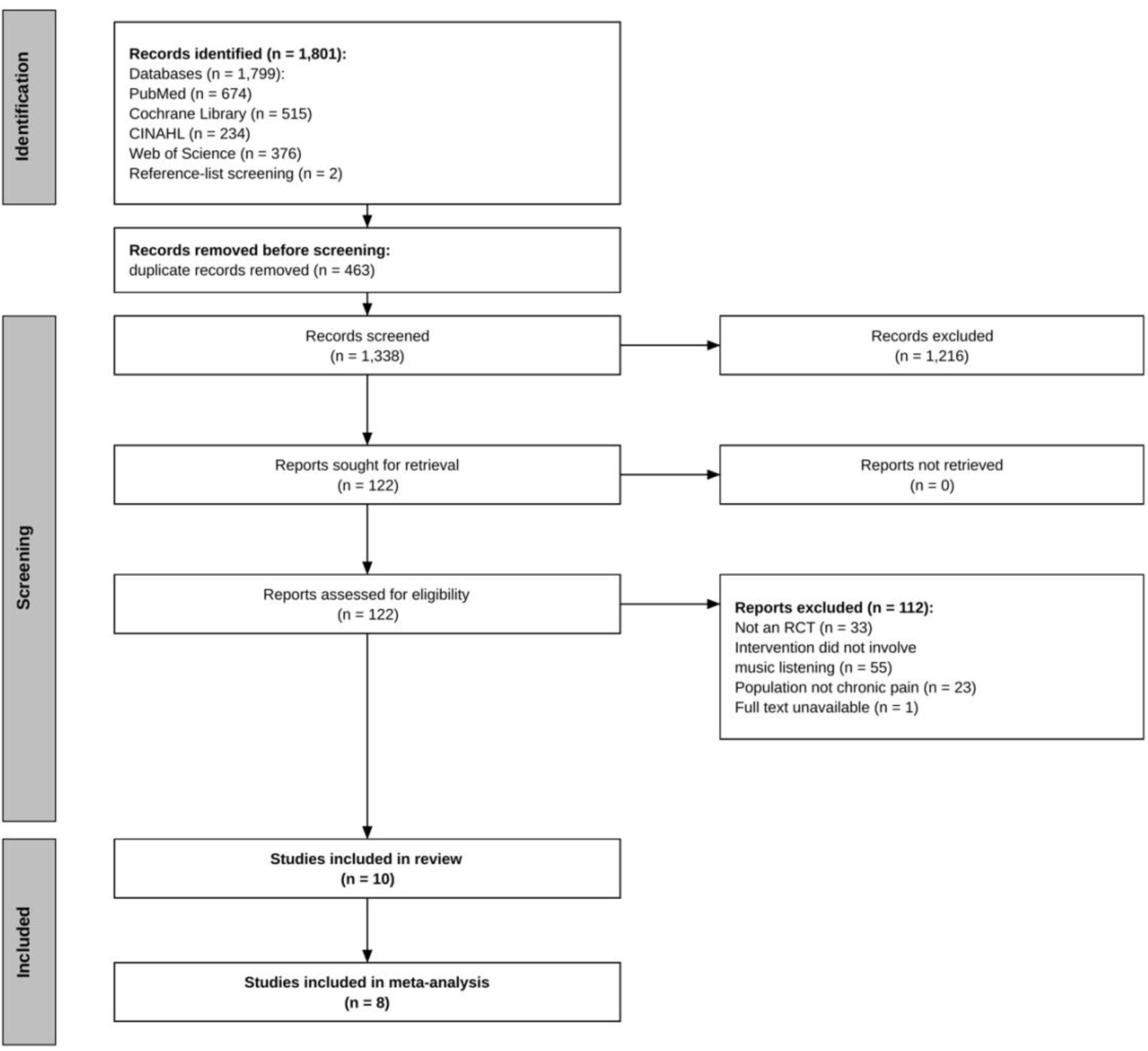
**PRISMA flow diagram of the study selection process.**

### 3.2. Study characteristics

The characteristics of the included studies are summarized in **Table 1** and described below according to study design, participant characteristics, intervention protocols, and outcomes.

**Table 1.**
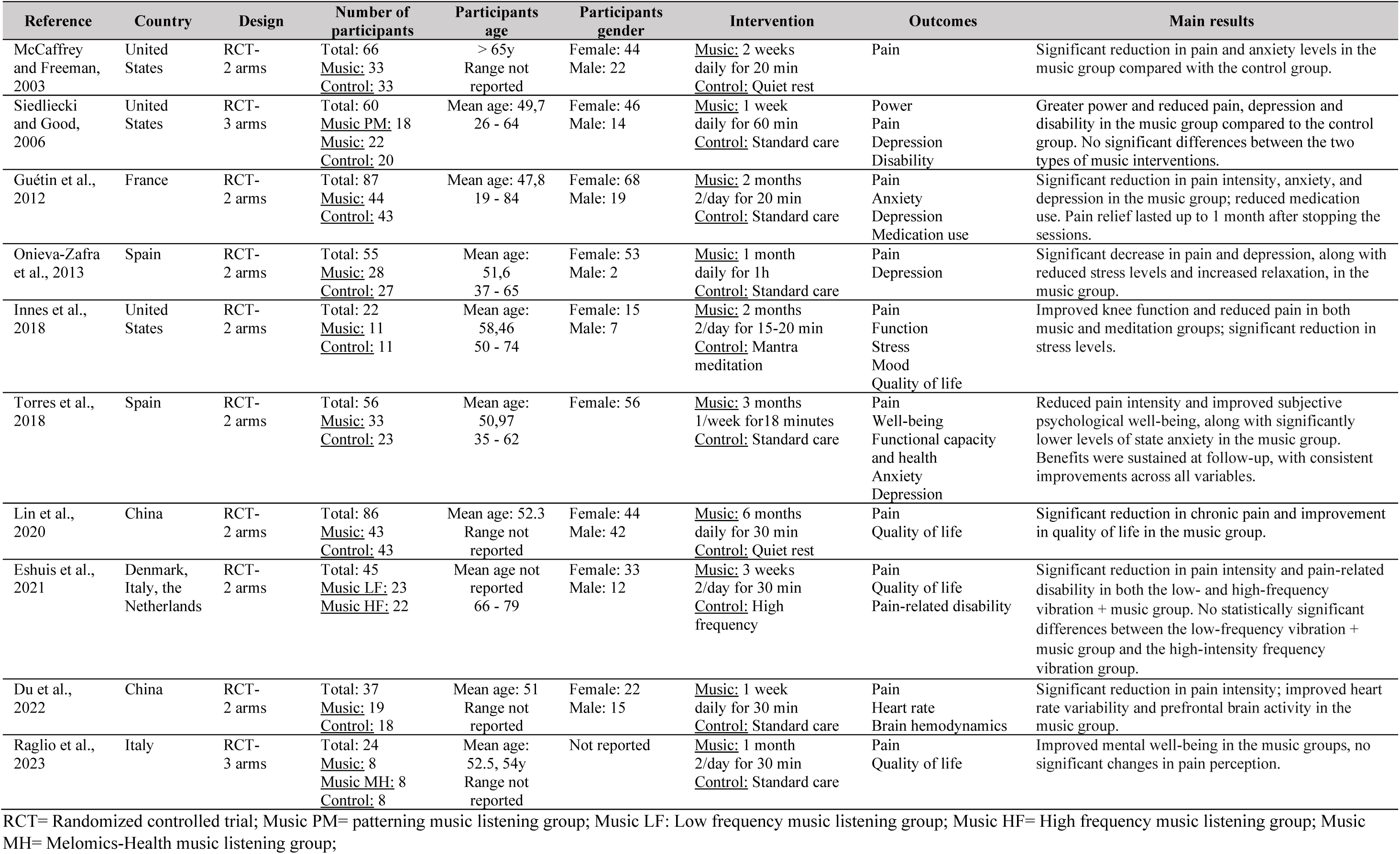
Characteristics of included studies.

| Reference | Country | Design | Number of participants | Participants age | Participants gender | Intervention | Outcomes | Main results |
| --- | --- | --- | --- | --- | --- | --- | --- | --- |
| McCaffrey and Freeman, 2003 | United States | RCT-2 arms | Total: 66<br><u>Music</u>: 33<br><u>Control</u>: 33 | > 65y<br>Range not reported | Female: 44<br>Male: 22 | <u>Music</u>: 2 weeks daily for 20 min<br><u>Control</u>: Quiet rest | Pain | Significant reduction in pain and anxiety levels in the music group compared with the control group. |
| Siedliecki and Good, 2006 | United States | RCT-3 arms | Total: 60<br><u>Music PM</u>: 18<br><u>Music</u>: 22<br><u>Control</u>: 20 | Mean age: 49,7<br>26 - 64 | Female: 46<br>Male: 14 | <u>Music</u>: 1 week daily for 60 min<br><u>Control</u>: Standard care | Power<br>Pain<br>Depression<br>Disability | Greater power and reduced pain, depression and disability in the music group compared to the control group. No significant differences between the two types of music interventions. |
| Guétin et al., 2012 | France | RCT-2 arms | Total: 87<br><u>Music</u>: 44<br><u>Control</u>: 43 | Mean age: 47,8<br>19 - 84 | Female: 68<br>Male: 19 | <u>Music</u>: 2 months 2/day for 20 min<br><u>Control</u>: Standard care | Pain<br>Anxiety<br>Depression<br>Medication use | Significant reduction in pain intensity, anxiety, and depression in the music group; reduced medication use. Pain relief lasted up to 1 month after stopping the sessions. |
| Onieva-Zafra et al., 2013 | Spain | RCT-2 arms | Total: 55<br><u>Music</u>: 28<br><u>Control</u>: 27 | Mean age: 51,6<br>37 - 65 | Female: 53<br>Male: 2 | <u>Music</u>: 1 month daily for 1h<br><u>Control</u>: Standard care | Pain<br>Depression | Significant decrease in pain and depression, along with reduced stress levels and increased relaxation, in the music group. |
| Innes et al., 2018 | United States | RCT-2 arms | Total: 22<br><u>Music</u>: 11<br><u>Control</u>: 11 | Mean age: 58,46<br>50 - 74 | Female: 15<br>Male: 7 | <u>Music</u>: 2 months 2/day for 15-20 min<br><u>Control</u>: Mantra meditation | Pain<br>Function<br>Stress<br>Mood<br>Quality of life | Improved knee function and reduced pain in both music and meditation groups; significant reduction in stress levels. |
| Torres et al., 2018 | Spain | RCT-2 arms | Total: 56<br><u>Music</u>: 33<br><u>Control</u>: 23 | Mean age: 50,97<br>35 - 62 | Female: 56 | <u>Music</u>: 3 months 1/week for 18 minutes<br><u>Control</u>: Standard care | Pain<br>Well-being<br>Functional capacity and health<br>Anxiety<br>Depression | Reduced pain intensity and improved subjective psychological well-being, along with significantly lower levels of state anxiety in the music group. Benefits were sustained at follow-up, with consistent improvements across all variables. |
| Lin et al., 2020 | China | RCT-2 arms | Total: 86<br><u>Music</u>: 43<br><u>Control</u>: 43 | Mean age: 52.3<br>Range not reported | Female: 44<br>Male: 42 | <u>Music</u>: 6 months daily for 30 min<br><u>Control</u>: Quiet rest | Pain<br>Quality of life | Significant reduction in chronic pain and improvement in quality of life in the music group. |
| Eshuis et al., 2021 | Denmark, Italy, the Netherlands | RCT-2 arms | Total: 45<br><u>Music LF</u>: 23<br><u>Music HF</u>: 22 | Mean age not reported<br>66 - 79 | Female: 33<br>Male: 12 | <u>Music</u>: 3 weeks 2/day for 30 min<br><u>Control</u>: High frequency | Pain<br>Quality of life<br>Pain-related disability | Significant reduction in pain intensity and pain-related disability in both the low- and high-frequency vibration + music group. No statistically significant differences between the low-frequency vibration + music group and the high-intensity frequency vibration group. |
| Du et al., 2022 | China | RCT-2 arms | Total: 37<br><u>Music</u>: 19<br><u>Control</u>: 18 | Mean age: 51<br>Range not reported | Female: 22<br>Male: 15 | <u>Music</u>: 1 week daily for 30 min<br><u>Control</u>: Standard care | Pain<br>Heart rate<br>Brain hemodynamics | Significant reduction in pain intensity; improved heart rate variability and prefrontal brain activity in the music group. |
| Raglio et al., 2023 | Italy | RCT-3 arms | Total: 24<br><u>Music</u>: 8<br><u>Music MH</u>: 8<br><u>Control</u>: 8 | Mean age: 52.5, 54y<br>Range not reported | Not reported | <u>Music</u>: 1 month 2/day for 30 min<br><u>Control</u>: Standard care | Pain<br>Quality of life | Improved mental well-being in the music groups, no significant changes in pain perception. |

RCT= Randomized controlled trial; Music PM= patterning music listening group; Music LF: Low frequency music listening group; Music HF= High frequency music listening group; Music MH= Melomics-Health music listening group;

#### Study identification details and design

The 10 included randomized controlled trials were published between 2003 and 2023, all in English. The studies were conducted across seven countries: three in the United States (Innes et al., 2018; McCaffrey & Freeman, 2003; Siedliecki & Good, 2006), two in China (Du et al., 2022a; Lin et al., 2020), two in Spain (Onieva-Zafra et al., 2013; Torres et al., 2018), one in Italy (Raglio et al., 2023), one in France (Guétin et al., 2012), and one multicenter study conducted across Denmark, Italy, and the Netherlands (Eshuis et al., 2021). The trials were published in journals spanning diverse disciplines, including pain, neuroscience, neurology, psychology, nursing, music therapy, surgery, and complementary medicine. Eight studies used a two-arm parallel-group design, whereas two included three intervention groups (Raglio et al., 2023; Siedliecki & Good, 2006).

#### Participant characteristics

The 10 included trials enrolled a total of 538 participants, with sample sizes ranging from 22 to 87 individuals. Across the included studies, participants were predominantly women (74.2%), with reported ages ranging from 26 to 84 years. Although most studies included comparable numbers of participants in the intervention and control groups, one trial allocated more participants to the music intervention than to the control condition (Torres et al., 2018).

Most studies investigated adults with chronic musculoskeletal pain (Eshuis et al., 2021; Guétin et al., 2012; Innes et al., 2018; McCaffrey & Freeman, 2003; Onieva-Zafra et al., 2013; Raglio et al., 2023; Torres et al., 2018), including fibromyalgia, osteoarthritis, chronic neuropathic pain, and non-specific musculoskeletal pain. The remaining studies evaluated participants with chronic non-cancer pain (Du et al., 2022a) (Siedliecki & Good, 2006), or chronic post-surgical pain following mechanical valve replacement (Lin et al., 2020), indicating that music listening has been investigated across a broad spectrum of chronic pain conditions.

Eligibility criteria were generally well defined and largely comparable across studies. Chronic pain was consistently defined as pain persisting for more than three months, and most trials required participants to report at least moderate pain intensity. Common exclusion criteria included severe psychiatric or cognitive disorders, hearing impairment, epilepsy, substance abuse, and previous participation in structured music-based interventions.

#### Intervention setting

Music listening interventions were delivered in both clinical and home settings. Two studies implemented the intervention under supervised clinical conditions with controlled acoustic environments (Eshuis et al., 2021; Torres et al., 2018), whereas most employed self-administered home-based interventions, allowing participants to integrate music listening into their daily routines (Du et al., 2022a; Guétin et al., 2012; Innes et al., 2018; Lin et al., 2020; McCaffrey & Freeman, 2003; Onieva-Zafra et al., 2013; Raglio et al., 2023). One study did not explicitly report the intervention setting (Siedliecki & Good, 2006).

#### Intervention characteristics

The main characteristics of the interventions are summarized in **Table 2**. Intervention protocols varied substantially in duration, frequency, session length, music selection procedures, and comparator conditions. Treatment duration ranged from one week (Du et al., 2022a; Siedliecki & Good, 2006) to six months (Lin et al., 2020), although most lasted between one and three months. Daily listening sessions were the most common approach, while several studies implemented two sessions per day (Eshuis et al., 2021; Guétin et al., 2012; Innes et al., 2018; Raglio et al., 2023). Session duration ranged from 6 to 60 minutes, with the exception of the multicomponent intervention by (Torres et al., 2018), which was delivered weekly over two-hour sessions but included only 6–18 minutes of music listening.

**Table 2.**
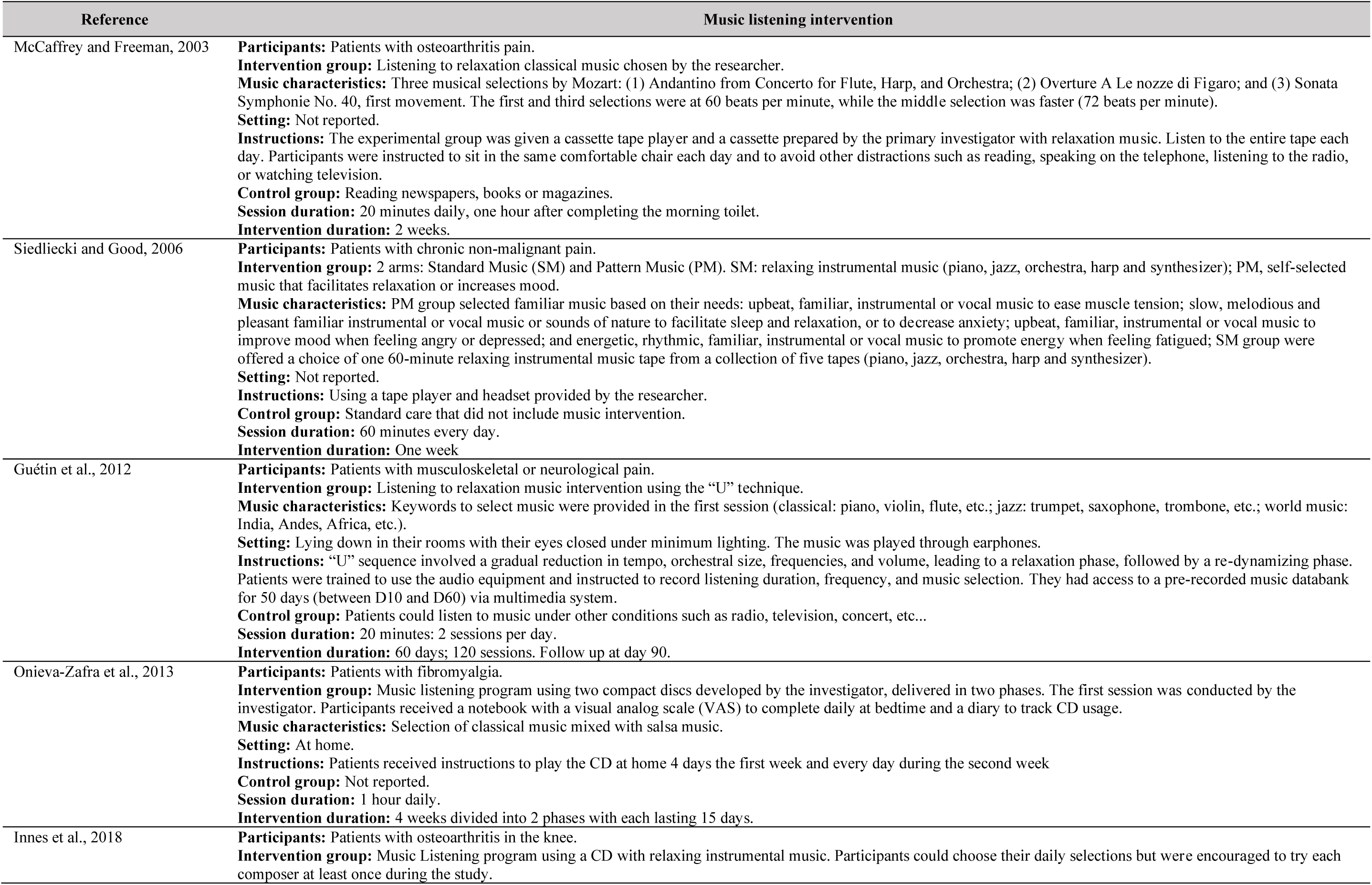
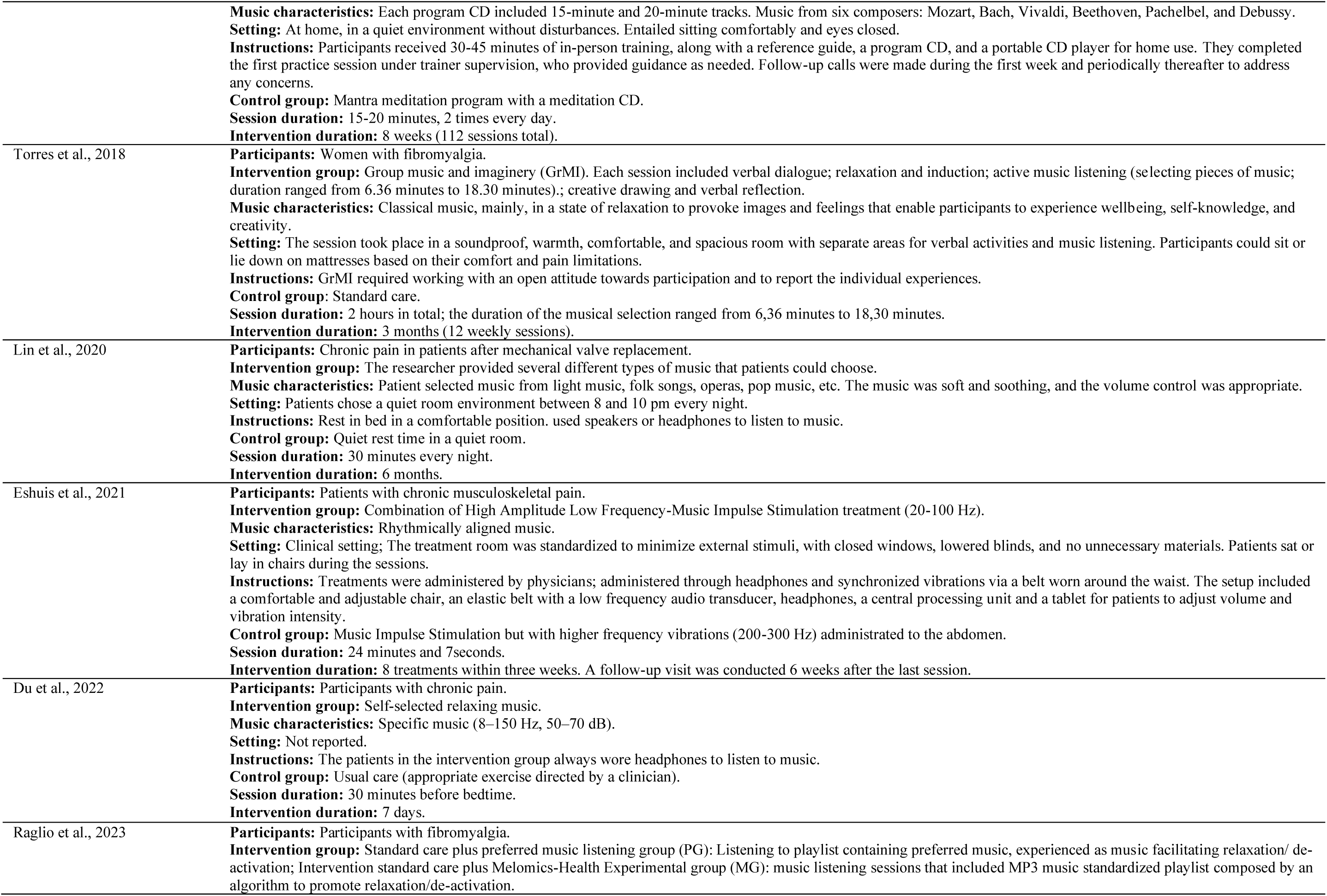
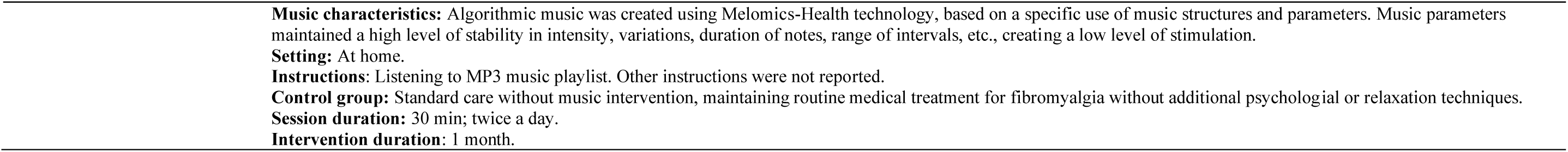
Main characteristics of the interventions.

Music selection procedures also varied across studies. Most interventions allowed participants to choose from playlists curated by the researchers (Eshuis et al., 2021; Guétin et al., 2012; Innes et al., 2018; Lin et al., 2020; McCaffrey & Freeman, 2003; Onieva-Zafra et al., 2013; Siedliecki & Good, 2006; Torres et al., 2018), whereas two studies employed self-selected music (Du et al., 2022a; Raglio et al., 2023). The music used across studies included relaxing, classical, instrumental, popular, folk, opera, and salsa music. Some interventions were designed around specific acoustic properties rather than musical genre. For example, two studies employed low-frequency music (16-50 Hz), intended to promote sleep and mood stabilization (Du et al., 2022a; McCaffrey & Freeman, 2003), whereas one trial combined high-amplitude, low-frequency music with impulse stimulation delivered through a dedicated vibroacoustic belt worn around the waist (Eshuis et al., 2021). Guided imagery was incorporated alongside music listening in one multicomponent intervention (Torres et al., 2018). Music was consistently delivered through headphones using audio playback devices, ranging from cassette tapes and compact discs in earlier studies to digital playlists and multimedia devices in more recent trials.

Only four studies reported monitoring treatment adherence. They used diaries, mobile applications, or multimedia software to document participants’ engagement (Du et al., 2022b; Guétin et al., 2012; Innes et al., 2018; Raglio et al., 2023). Similarly, only three studies described modifications to the intervention based on participants’ responses or tolerance (Eshuis et al., 2021; Guétin et al., 2012; Torres et al., 2018), highlighting the principal reporting deficiencies identified across the included studies.

Comparator conditions were heterogeneous. Six studies used usual care as the control condition (Du et al., 2022a; Guétin et al., 2012; Onieva-Zafra et al., 2013; Raglio et al., 2023; Siedliecki & Good, 2006; Torres et al., 2018), whereas the remaining trials employed alternative comparators, including vibroacoustic stimulation with higher-frequency vibrations (Eshuis et al., 2021), quiet rest (Lin et al., 2020), guided imagery (Torres et al., 2018), or mantra meditation (Innes et al., 2018).

### 3.3. Quality of intervention reporting

Application of the TIDieR checklist showed that the overall completeness of intervention reporting was high, although several studies omitted key information required for replication (**Table 3**). All trials provided a clear description of the intervention rationale, materials, procedures, mode of delivery, setting, dose, and tailoring, allowing the interventions to be understood and potentially replicated. However, reporting of intervention modifications made in response to participant feedback was limited, and monitoring of treatment adherence and intervention fidelity was reported in only a minority of studies. These were the only TIDieR items that were consistently underreported across the included trials, representing the principal reporting deficiencies identified in the current evidence base.

**Table 3.** Studies and items of the TIDieR checklist.

| TIDieR Checklist Item | McCaffrey and Freeman, 2003 | Siedliecki and Good, 2006 | Guétin et al., 2012 | Onieva-Zafra et al., 2013 | Innes et al., 2018 | Torres et al., 2018 | Lin et al., 2020 | Eshuis et al., 2021 | Du et al., 2022 | Raglio et al., 2023 |
| --- | --- | --- | --- | --- | --- | --- | --- | --- | --- | --- |
| 1. *Brief name* | ✓ | ✓ | ✓ | ✓ | ✓ | ✓ | ✓ | ✓ | ✓ | ✓ |
| 2. *Why (rationale)* | ✓ | ✓ | ✓ | ✓ | ✓ | ✓ | ✓ | ✓ | ✓ | ✓ |
| 3. *Materials used in intervention:* | ✓ | ✓ | ✓ | ✓ | ✓ | ✓ | ✓ | ✓ | ✓ | ✓ |
| 4. *Procedures:* | ✓ | ✓ | ✓ | ✓ | ✓ | ✓ | ✓ | ✓ | ✓ | ✓ |
| 5. *Intervention Providers:* | ✓ | ✓ | ✓ | ✓ | ✓ | ✓ | ✓ | ✓ | ✓ | ✓ |
| 6. *Modes of delivery* | ✓ | ✓ | ✓ | ✓ | ✓ | ✓ | ✓ | ✓ | ✓ | ✓ |
| 7. *Location of the intervention.* | ✗ | ✓ | ✓ | ✓ | ✓ | ✓ | ✓ | ✓ | ✗ | ✓ |
| 8. *When and how much:* | ✓ | ✓ | ✓ | ✓ | ✓ | ✓ | ✓ | ✓ | ✓ | ✓ |
| 9. *Details of Tailoring:* | ✓ | ✓ | ✓ | ✓ | ✓ | ✓ | ✓ | ✓ | ✓ | ✓ |
| 10. *Modifications to intervention* | ✗ | ✗ | ✓ | ✗ | ✗ | ✓ | ✗ | ✓ | ✗ | ✗ |
| 11. *How well (fidelity, adherence)* | ✗ | ✗ | ✓ | ✗ | ✓ | ✗ | ✗ | ✗ | ✓ | ✓ |

TIDieR: Template for Intervention Description and Replication checklist. ✓= item adequately reported; ✗= item inadequately or not reported

### 3.4. Risk of bias in included studies

**Figure 2** summarizes the risk of bias assessment across the included trials.

**Figure 2.**
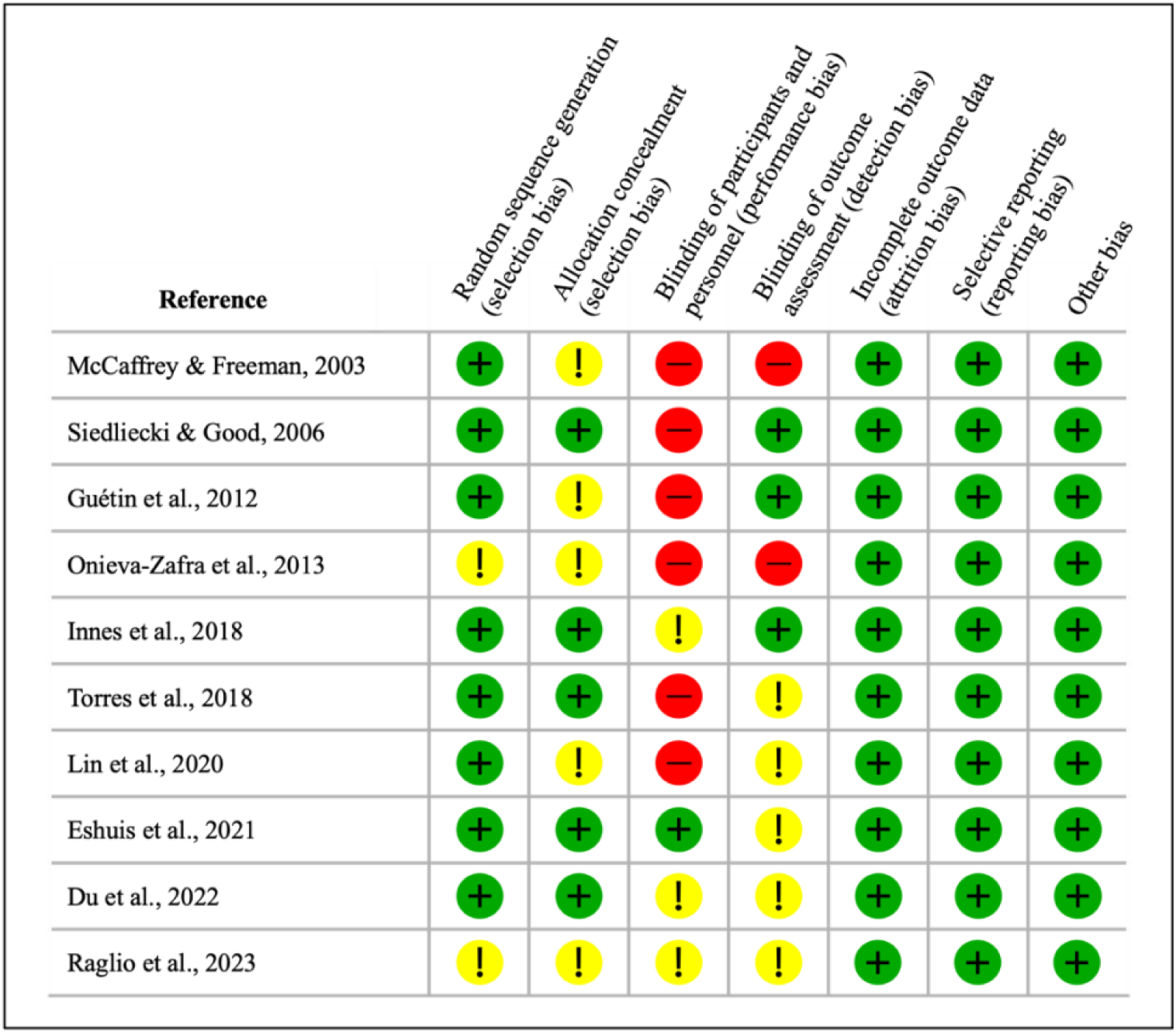
Risk of bias in included studies. Note. Summary of the risk of bias assessment for each included trial according to the Cochrane Risk of Bias Tool. Green circles indicate low risk of bias, yellow circles indicate unclear risk, and red circles indicate high risk.

#### Random sequence generation

Eight studies reported the randomization method used to generate the randomization sequence, including computer-generated sequences or marked slips of paper, and were judged to have a low risk of selection bias (Du et al., 2022a; Eshuis et al., 2021; Guétin et al., 2012; Innes et al., 2018; Lin et al., 2020; McCaffrey & Freeman, 2003; Siedliecki & Good, 2006; Torres et al., 2018). Two studies did not provide sufficient information on sequence generation and were therefore rated as having an unclear risk of bias (Onieva-Zafra et al., 2013; Raglio et al., 2023).

#### Allocation concealment

Allocation concealment methods were adequately reported in five studies (Du et al., 2022a; Eshuis et al., 2021; Innes et al., 2018; Siedliecki & Good, 2006; Torres et al., 2018), whereas the remaining trials did not describe the concealment procedure sufficiently to permit judgement (Guétin et al., 2012; Lin et al., 2020; McCaffrey & Freeman, 2003; Onieva-Zafra et al., 2013; Raglio et al., 2023).

#### Blinding of participants and personnel

Blinding of participants and personnel represented the domain at greatest risk of bias. In six studies, neither participants nor study personnel were blinded (Guétin et al., 2012; Lin et al., 2020; McCaffrey & Freeman, 2003; Onieva-Zafra et al., 2013; Siedliecki & Good, 2006; Torres et al., 2018), while three studies provided insufficient information to judge the risk of bias (Innes et al., 2018; Du et al., 2022; Raglio et al., 2023). Only one successfully implemented participant blinding by using identical equipment delivering music at different frequencies (Eshuis et al., 2021).

#### Blinding of outcome assessment

Outcome assessor blinding was explicitly reported in only two studies (Guétin et al., 2012; Innes et al., 2018). The remaining trials did not provide sufficient information to determine whether outcome assessors were blinded, resulting in an unclear risk of detection bias.

#### Incomplete outcome data

Risk of attrition bias was generally low. All studies retained more than 85% of participants and appropriately addressed incomplete outcome data in their analyses.

#### Selective reporting

All included studies were judged to have a low risk of selective reporting, as the reported outcomes were consistent with those specified in the study methods or trial protocols when available.

#### Other bias

No important additional sources of bias, including baseline imbalances, conflicts of interest, or funding-related concerns, were identified across the included studies.

### 3.5. Effects of music listening interventions

Across the included trials, music listening was consistently associated with improvements in pain and several psychological and functional outcomes. This section qualitatively summarizes the effects of music listening on pain, anxiety, depression, mood, quality of life, and functionality. The outcome measures used across studies are summarized in **Table 4**, whereas the quantitative synthesis of pain intensity is presented separately in Section 3.6.

**Table 4.** Outcome measures and their corresponding scales.

| **Outcome** | **Standardized assessment** |
| --- | --- |
| **Pain** | Visual Analog Scale (VAS)<br>McGill Pain Questionnaire<br>Pain Intensity and Impact brief pain inventory (BPI-PS)<br>Knee Injury and Osteoarthritis Outcome Score (KOOS)<br>Pain Disability Index (PDI)<br>Pain Catastrophizing Scale (PCS)<br>Numeric Rating Scale (NRS) |
| **Anxiety** | Hospital Anxiety and Depression Scale (HADS)<br>State and Trait Anxiety Levels |
| **Depression** | Beck Depression Inventory (BDI)<br>State-Trait Depression Questionnaire |
| **Stress** | Perceived Stress Scale |
| **Mood** | Profile of Mood States (POMS)<br>Cognitive Behavioral Assessment Outcome Evaluation (CBA-OE) |
| **Wellbeing** | Psychological Well-Being Scale (PWBS) |
| **Disability** | Pain Disability Index (PDI) |
| **Functionality** | Knee Injury and Osteoarthritis Outcome Score (KOOS)<br>Fibromyalgia Impact Questionnaire (FIQ) |

#### Pain

Pain was assessed using a range of validated instruments, including the Visual Analog Scale (VAS), Numeric Rating Scale (NRS), McGill Pain Questionnaire (MPQ), Brief Pain Inventory (BPI-PS), Knee Injury and Osteoarthritis Outcome Score (KOOS), Pain Disability Index (PDI), and Pain Catastrophizing Scale (PCS) (Table 4).

All included studies evaluated pain outcomes. Overall, music listening was consistently associated with reductions in pain intensity and pain-related outcomes, with only one trial reporting no significant improvement in pain intensity (Raglio et al., 2023). Several studies demonstrated clinically meaningful reductions in pain following interventions based on relaxing, classical, or low-frequency music (Du et al., 2022a; Eshuis et al., 2021; Guétin et al., 2012; Innes et al., 2018; Lin et al., 2020; McCaffrey & Freeman, 2003; Siedliecki & Good, 2006; Torres et al., 2018), while interventions incorporating different musical genres, including classical music and salsa, also produced significant analgesic effects (Onieva-Zafra et al., 2013). Notably, (Guétin et al., 2012) reported that pain reduction persisted for up to one month after the intervention. No consistent differences in effectiveness were observed between interventions using self-selected music and those employing researcher-selected playlists.

#### Anxiety and depression

Anxiety and depression were evaluated in four studies (Guétin et al., 2012; Onieva-Zafra et al., 2013; Siedliecki & Good, 2006; Torres et al., 2018), while two studies additionally assessed perceived stress (Innes et al., 2018; Onieva-Zafra et al., 2013). Psychological outcomes were measured using validated instruments, including the Hospital Anxiety and Depression Scale (HADS), Beck Depression Inventory (BDI), State-Trait Anxiety Inventory (STAI), State-Trait Depression Questionnaire (ST-DEP), and Perceived Stress Scale (PSS) (Table 4).

Overall, music listening was consistently associated with improvements in psychological wellbeing. Most studies reported significant reductions in anxiety and depressive symptoms following the intervention (Guétin et al., 2012; Onieva-Zafra et al., 2013; Siedliecki & Good, 2006; Torres et al., 2018), and one study also documented a reduced use of anxiolytic medication among participants receiving music listening (Guétin et al., 2012). Although (Du et al., 2022a) did not directly assess psychological outcomes, improvements in heart rate variability suggested a reduction in physiological stress. Across the studies evaluating depression, beneficial effects were observed irrespective of whether participants listened to self-selected or researcher-selected music, including interventions based on relaxing music and combinations of classical and salsa music.

#### Mood and psychological well-being

Mood and psychological well-being were evaluated in three studies using the Profile of Mood States (POMS), the Psychological Well-Being Scale (PWBS), and the Cognitive Behavioural Assessment Outcome Evaluation (CBA-OE) **(Table 4**) (Innes et al., 2018; Raglio et al., 2023; Torres et al., 2018).

Overall, music listening was associated with improvements in mood and psychological well-being. (Innes et al., 2018) reported improved mood and reduced perceived stress following self-selected relaxing instrumental music, although similar benefits were also observed in the active control group receiving mantra meditation. (Raglio et al., 2023) found improvements in well-being among patients with fibromyalgia listening to both preferred and algorithmically selected music, with only the algorithmic music group maintaining these benefits at follow-up. Similarly, (Torres et al., 2018) reported significant improvements in subjective psychological well-being following a multicomponent intervention combining guided relaxation and music listening.

#### Quality of life

Quality of life was evaluated in four studies using the quality-of-life subscale of the Knee Injury and Osteoarthritis Outcome Score (KOOS), the Short Form-36 Health Survey (SF-36), the Short Form-12 Health Survey (SF-12), and the EuroQol 5-Dimension 3-Level questionnaire (EQ-5D-3L) (**Table 4**) (Eshuis et al., 2021; Innes et al., 2018; Lin et al., 2020; Raglio et al., 2023).

Overall, music listening was associated with improvements in quality of life across different chronic pain populations. Positive effects were reported in three of the four studies (Innes et al., 2018; Lin et al., 2020; Raglio et al., 2023), whereas (Eshuis et al., 2021), found no significant changes following an intervention combining music listening with low-frequency vibroacoustic stimulation.

#### Functionality in daily activities

Functionality and disability were evaluated in four studies using the Pain Disability Index (PDI), the Knee Injury and Osteoarthritis Outcome Score (KOOS), and the Fibromyalgia Impact Questionnaire (FIQ) (**Table 4**) (Eshuis et al., 2021; Innes et al., 2018; Siedliecki & Good, 2006; Torres et al., 2018).

Findings regarding functional outcomes were less consistent than those observed for pain and psychological wellbeing. Two studies reported significant improvements in disability and functional capacity following music listening interventions (Siedliecki & Good, 2006; Torres et al., 2018), whereas no functional benefit was observed in the trial combining music listening with low-frequency vibroacoustic stimulation (Eshuis et al., 2021). In older adults with osteoarthritis, improvements in knee function and sports-related activity were greater following an active mantra meditation programme than after passive music listening (Innes et al., 2018).

### 3.6. Meta-analysis of pain outcomes

Eight randomized controlled trials, contributing ten effect estimates, were included in the meta-analysis of pain intensity. The pooled analysis demonstrated a significant benefit of music listening compared with control conditions, with an SMD of −0.53 (95% CI: −0.96 to −0.11; *p* = 0.014), corresponding to a moderate reduction in pain intensity. Considerable between-study heterogeneity was observed (Q(9) = 35.80, *p* < 0.001; I² = 76.16%).

Assessment of publication bias did not suggest the presence of small-study effects. Egger’s regression test (*p* = 0.209) and Kendall’s tau (*p* = 0.156) were both non-significant, visual inspection of the funnel plot did not indicate substantial asymmetry, and the fail-safe N showed that 94 additional null studies would be required to render the pooled effect non-significant.

The forest plot summarizing the meta-analysis is presented in **Figure 3**.

**Figure 3.**
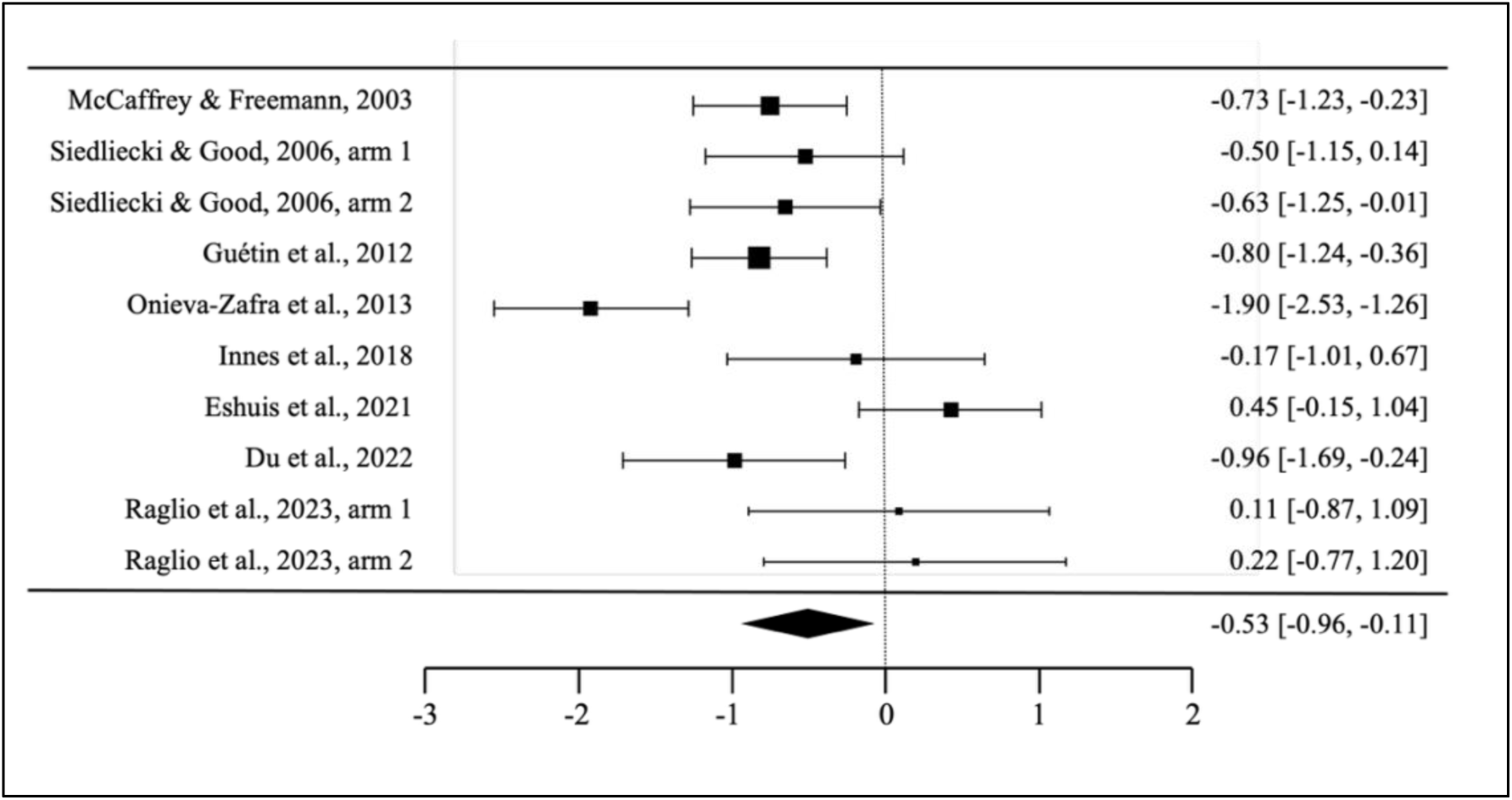
Pooled standardized mean differences for pain intensity outcomes comparing music listening and control conditions. Note. Each horizontal line represents an individual study’s standardized mean difference (SMD) and 95% confidence interval (CI). Negative values indicate lower pain intensity in the music listening group compared with control conditions. The diamond represents the pooled effect estimate (SMD = −0.53, 95% CI: −0.96 to −0.11), showing a significant moderate reduction in pain following music listening, with high heterogeneity (I² = 76.16%). In Siedliecki & Good (2006), arm 1 corresponds to the patterned music group, which received rhythmically structured music sessions for relaxation and distraction, whereas arm 2 corresponds to the standard music group, who listened to ambient music for comfort. In Raglio et al. (2023), arm 1 corresponds to the playlist group, who listened to preferred pre-selected music, and arm 2 to the Melomics group, who received algorithmically generated music designed to modulate emotional and physiological responses.

## 4. Discussion

This systematic review and meta-analysis confirms that music listening is an effective complementary intervention for chronic pain management, providing benefits that extend beyond pain reduction to psychological wellbeing, quality of life, and functional outcomes. Although the meta-analysis demonstrated a moderate analgesic effect, substantial between-study heterogeneity remained. Our findings suggest that the central challenge for the field has shifted from establishing whether music listening is effective to determining how effective interventions can be reproduced and implemented consistently across clinical settings. Achieving this goal will require more rigorous and transparent reporting of intervention protocols.

### 4.1. Pain

The present findings provide consistent evidence that music listening reduces pain intensity in individuals with chronic pain. Beyond reductions in pain intensity measured with validated instruments such as the Visual Analogue Scale (VAS), Numerical Rating Scale (NRS), and Brief Pain Inventory (BPI), several studies also reported benefits in broader dimensions of the pain experience, including disability and pain catastrophizing. These findings reinforce the view that music listening influences not only the sensory component of pain but also its cognitive and affective dimensions.

The meta-analysis confirmed a moderate analgesic effect, although between-study heterogeneity was considerable. This variability likely reflects differences in patient populations, pain conditions, intervention characteristics, and adherence. It may also indicate that the therapeutic effects of music depend on multiple interacting components rather than on a single intervention feature. One of the most debated questions concerns whether music should be selected by participants or prescribed by researchers. Although previous reviews have suggested advantages of participant-selected music (Garza-Villarreal et al., 2017; Hsu et al., 2022), the evidence synthesized here does not support clear superiority of either approach. Instead, both strategies appear capable of producing meaningful clinical benefits when appropriately implemented.

Similarly, while slow-tempo, relaxing, instrumental, or classical music was most frequently associated with positive outcomes, the interpretation of these findings is constrained by incomplete reporting of musical characteristics. Interventions are often described simply as “relaxing music”, with little information on tempo, rhythmic structure, harmonic content, or other relevant musical features. Consequently, it remains difficult to determine which musical components are responsible for the observed analgesic effects. More comprehensive reporting of intervention characteristics will be essential for identifying the active ingredients of music listening and for developing reproducible, evidence-based clinical protocols.

### 4.2. Psychological wellbeing, quality of life, and functionality

The benefits of music listening extended beyond analgesia, reinforcing the view that chronic pain should be approached as a multidimensional condition rather than a purely sensory experience. Across the included studies, music listening was associated with reductions in anxiety and depression, improvements in mood, enhanced quality of life, and, where evaluated, better functional outcomes. These findings suggest that music may simultaneously target several of the interconnected dimensions that sustain chronic pain.

The mechanisms underlying these broader benefits are likely multifactorial. Music influences limbic and paralimbic networks involved in emotional regulation, including the amygdala and anterior cingulate cortex, while promoting physiological relaxation through reductions in heart rate, respiration, muscle tension, and stress-related hormonal responses (Arnold et al., 2024; Harney et al., 2023; Mallik & Russo, 2022). Together with cognitive mechanisms such as distraction and positive emotional engagement, these effects may improve coping, reduce emotional distress, and increase resilience, illustrating the interplay of psychological, physiological, and neurobiological mechanisms underlying music-induced analgesia (Arnold et al., 2024; Fernández-Dueñas et al., 2026).

Improvements in quality of life and functionality are likely secondary to these combined effects. By reducing pain and psychological distress, music listening may facilitate participation in everyday activities, increase social engagement, and encourage physical movement, helping individuals regain a sense of normality and control over their lives (Arnold et al., 2024; Pothoulaki et al., 2008; Thoma et al., 2012). Although functionality was assessed in relatively few studies, the available evidence suggests that music may indirectly improve functional outcomes by addressing barriers such as pain-related fear, anxiety, and depressive symptoms (Innes et al., 2018; Siedliecki & Good, 2006; Torres et al., 2018). These findings further support the role of music listening as an adjunctive intervention capable of addressing the broader biopsychosocial consequences of chronic pain, rather than pain intensity alone.

### 4.3. Intervention characteristics and implementation considerations

A central finding of this review is the marked heterogeneity of music listening interventions. Protocols differed considerably in setting, duration, frequency, supervision, music selection procedures, and comparator conditions. Despite this variability, beneficial effects were observed across a diverse delivery formats, highlighting the flexibility and scalability of music listening as a complementary intervention for chronic pain. At the same time, this diversity likely contributed to the substantial heterogeneity observed in the meta-analysis and limits the identification of the intervention characteristics that are most strongly associated with clinical benefit.

The available evidence does not allow firm recommendations regarding the optimal intervention protocol. Nevertheless, interventions involving regular listening sessions of approximately 30 minutes or longer and those maintained over several weeks appeared more likely to produce sustained improvements. Similarly, both participant-selected and researcher-selected music were associated with positive outcomes, suggesting that successful implementation may depend less on how music is selected than on delivering a structured and engaging intervention.

The considerable variability in intervention design should therefore be viewed not simply as a methodological limitation but as a major barrier to clinical translation. Until music listening protocols are described more consistently and key intervention components are reported transparently, it will remain difficult to identify the active ingredients of treatment, compare findings across studies, and establish evidence-based recommendations for routine clinical practice.

### 4.4. Quality of intervention reporting and risk of bias

A major strength of this review is that, beyond synthesizing the evidence on effectiveness, it systematically evaluated the completeness of intervention reporting using the TIDieR framework. Overall, the included trials adequately described the general structure of their interventions, including the rationale, materials, delivery procedures, setting, and treatment dose. However, two components were consistently underreported across studies: intervention modifications introduced during the trial and the assessment of treatment fidelity and participant adherence. These represent the most important and systematic reporting deficiencies identified in the current literature.

These omissions are not merely methodological shortcomings. Without information on adherence and treatment fidelity, it is impossible to determine whether observed outcomes reflect the intervention as designed or the extent to which participants actually engaged with it. Likewise, without documenting protocol modifications, successful adaptations cannot be replicated or implemented in future studies or routine clinical practice. For an intervention whose principal advantages are its accessibility, low cost, and scalability, transparent reporting of these components is essential for successful clinical translation. The routine use of reporting frameworks such as TIDieR during both study design and manuscript preparation should therefore become standard practice in future music listening trials.

Overall, the methodological quality of the included studies was satisfactory, with risk of bias generally rated as low. The most common limitation concerned the blinding of participants and study personnel, an inherent challenge in behavioural interventions requiring active participation. This limitation is common across non-pharmacological interventions and should be interpreted within the broader methodological context of complex intervention research rather than as a weakness specific to music listening.

### 4.5. Comparison with previous reviews

The present findings are consistent with previous systematic reviews and meta-analyses showing that music listening is associated with reductions in pain and improvements in psychological outcomes in individuals with chronic pain (Cepeda et al., 2006; Chen et al., 2025; Garza-Villarreal et al., 2017; Hsu et al., 2022; Lee, 2016; Martin-Saavedra et al., 2018). Recent evidence syntheses have similarly concluded that music provides benefits extending beyond pain intensity to emotional regulation, depression, anxiety, and other dimensions of wellbeing (Cournoyer Lemaire & Perreault, 2024) (Chen et al., 2025). However, discrepancies remain regarding the magnitude and consistency of these secondary outcomes, particularly for anxiety and quality of life. Rather than questioning the therapeutic potential of music listening, these inconsistencies likely reflect the considerable heterogeneity of intervention protocols and the incomplete reporting of key intervention components across studies.

The present review extends previous evidence in several important ways. By focusing exclusively on randomized controlled trials in chronic pain populations, it provides an updated quantitative estimate confirming a moderate analgesic effect of music listening. More importantly, it shifts the focus from the question of whether music listening is effective to whether effective interventions are described with sufficient completeness to be reproduced and implemented in clinical practice. To our knowledge, this is the first systematic review to combine an updated meta-analysis with a structured evaluation of intervention reporting quality using the TIDieR framework. Our findings suggest that future advances in the field will depend not only on generating additional efficacy data but also on improving the transparency, standardization, and reproducibility of intervention protocols.

### 4.6. Limitations and future directions

Several limitations should be considered when interpreting these findings. Although this review included all available randomized controlled trials, the evidence base remains relatively small and was characterized by considerable heterogeneity in participant populations, pain conditions, intervention protocols, comparator groups, and outcome measures. This variability limits the precision of the pooled effect estimate and precluded more detailed subgroup analyses to identify the intervention characteristics most strongly associated with clinical benefit. In addition, many studies reported limited information on treatment adherence, intervention fidelity, and long-term follow-up, restricting conclusions regarding the sustainability of treatment effects.

A further limitation concerns the characterization of the music itself. Interventions were frequently described using broad terms such as “relaxing music”, with little information on tempo, rhythmic structure, harmonic content, mode, or other acoustic properties that may influence therapeutic outcomes. Although the present review focused on the completeness of intervention reporting using the TIDieR framework, a more detailed characterization of musical features represents an important next step. Identifying which structural and perceptual components of music contribute most strongly to analgesic and psychological benefits will be essential for developing more precise and reproducible interventions, in line with current efforts to define the active ingredients of complex arts-based therapies (Grau-Sánchez et al., 2022b; Rodríguez-Fornells et al., 2025b; Skov & Nadal, 2025).

Future research should therefore move beyond simply demonstrating efficacy. Priority should be given to developing standardized intervention protocols, improving reporting transparency, incorporating rigorous monitoring of adherence and treatment fidelity, and extending follow-up assessments. At the same time, mechanistic studies integrating neuroimaging, psychophysiology, and behavioural outcomes may help clarify how specific intervention characteristics influence therapeutic response, ultimately supporting the development of evidence-based and clinically implementable music listening interventions for chronic pain.

## 5. Conclusions

This systematic review and meta-analysis demonstrates that music listening is a safe, accessible, and effective complementary intervention for chronic pain management. Beyond producing a moderate reduction in pain intensity, music listening improves psychological wellbeing, quality of life, and functional outcomes, reinforcing its value as an intervention that addresses the multidimensional nature of chronic pain. Importantly, these findings indicate that the principal challenge for the field is no longer establishing whether music listening is effective, but ensuring that effective interventions are described with sufficient completeness and consistency to be reproduced, compared across studies, and implemented in routine clinical practice. Improving the transparency and standardization of intervention reporting should therefore become a priority for future research. By facilitating the translation of evidence into practice, music listening has the potential to become an integral component of multidisciplinary chronic pain management and contribute to reducing reliance on long-term pharmacological therapies.

## 6. Declarations

### Ethics approval and consent to participate

Not applicable.

### Consent for publication

Not applicable.

### Availability of data and materials

Datasets used in the current study are available in the DDEUIT repository of the library of our university, https://euit.koha.es/cgi-bin/koha/opac-detail.pl?biblionumber=65531

### Authors’ contributions

**JGP** and **MTSV** conceptualized the study, performed the search, and evaluated the eligibility of articles according to the inclusion criteria. Both independently extracted data for subsequent comparison and combination, assessed the quality of intervention reporting and the risk of bias, and contributed to writing the original draft as well as to review and editing. **LZ, MFPT**, and **RV** conceptualized the study and contributed to writing, review, and editing. **ARF** and **VFD** conceptualized the study, contributed to the design and analysis, and to writing, review, and editing. **JGS** conceptualized the study and participated in all stages of the inclusion process, data extraction, and the evaluation of intervention reporting quality and risk of bias by resolving disagreements to reach consensus. She also designed and performed the analysis, and contributed to writing the original draft, review, and editing. All authors read and approved the final manuscript.

## Data Availability

All data analysed in this systematic review and meta-analysis were extracted from previously published studies, which are cited in the manuscript and reference list. All data produced in the present study are available upon reasonable request to the authors.

## Acknowledgements

We would like to thank Andreu Oliver Moreno for his valuable support with the analysis.

