## Supplementary Material for "Music listening for chronic pain management: a systematic review, meta-analysis, and evaluation of intervention reporting quality"

### S1. Search strategies

Systematic searches were conducted in four electronic databases from September 2004 to June 2025. A language filter was applied to include only articles published in English or Spanish. Reference lists of retrieved studies were hand-searched for additional records. The searches were run between September 2024 and June 2025.

#### PubMed (via MEDLINE)

("chronic pain" OR "pain management" OR "persistent pain")
AND ("music" OR "listening to music" OR "music induced analgesia" OR "musicotherapy" OR "music*")
AND (quality of life) OR (anxiety) OR (depression) OR (mood) OR (functionality) OR (activities of daily living) OR (daily living activities)

#### Cochrane Central Register of Controlled Trials (CENTRAL, via the Cochrane Library)

("chronic pain" OR "pain management" OR "persistent pain")
AND ("music" OR "listening to music" OR "music induced analgesia" OR "musicotherapy" OR "music*")
AND (quality of life) OR (anxiety) OR (depression) OR (mood) OR (functionality) OR (activities of daily living) OR (daily living activities)

#### CINAHL Complete (via EBSCOhost)

("chronic pain" OR "pain management" OR "persistent pain")
AND ("music" OR "listening to music" OR "music induced analgesia" OR "musicotherapy" OR "music*")
AND (quality of life) OR (anxiety) OR (depression) OR (mood) OR (functionality) OR (activities of daily living) OR (daily living activities)

#### Web of Science (via Clarivate)

("chronic pain" OR "pain management" OR "persistent pain")
AND ("music" OR "listening to music" OR "music induced analgesia" OR "musicotherapy" OR "music*")
AND (quality of life) OR (anxiety) OR (depression) OR (mood) OR (functionality) OR (activities of daily living) OR (daily living activities)

#### Records identified

PubMed: 674 · Cochrane Library: 515 · CINAHL: 234 · Web of Science: 376 · Reference-list screening: 2. Total identified: 1,799. After removing duplicates (n = 463), 1,338 records were screened by title/abstract; 122 full texts were assessed; 10 RCTs were included. See the PRISMA flow diagram (Figure 1) for the full selection process.

**S2. Eligibility criteria (PICOS)**
Studies were included or excluded according to the following pre-specified criteria, structured using the PICOS framework.

| **Criterion** | **Inclusion** | **Exclusion** |
|---|---|---|
| Population (P) | Adults aged ≥18 years with any type of chronic pain, defined as persistent or recurrent pain lasting longer than three months. | Participants with acute pain or oncological (cancer-related) pain. |
| Intervention (I) | Passive music listening delivered as a therapeutic intervention for chronic pain management. | Active music-based interventions (e.g., playing music, singing, or moving to music). |
| Comparison (C) | No restriction on the type of control intervention (e.g., usual/standard care, quiet rest, guided imagery, meditation, active comparators). | — |
| Outcomes (O) | Pain, plus one or more of: anxiety, depression, mood, quality of life, or functionality in activities of daily living. | Studies not reporting pain or any of the specified secondary outcomes. |
| Study design (S) | Randomized controlled trials (RCTs). | Non-randomized, quasi-experimental, or observational studies; reviews. |
| Other | Articles published in English or Spanish. No restriction on setting or publication date. | Articles published in languages other than English or Spanish. |

Selection process: Two reviewers (JGP and MTSV) independently screened titles/abstracts and then full texts. Disagreements at any stage were resolved by a third reviewer (JGS) to reach consensus. Duplicates were removed manually using Mendeley.

**S3. Data extraction sheet**

Data were extracted independently by two reviewers (JGP and MTSV) using a structured form; results were contrasted and combined, with disagreements resolved by a third reviewer (JGS). Where information was missing or unclear, study authors were contacted for clarification. The extraction fields are listed below, followed by the populated extraction table for the 10 included trials.

**Extraction fields**

**Study identification:**

- Authors, publication year, country, journal, study design

**Population characteristics:**

- Sample size (total; experimental and control groups), age range/mean, sex distribution, diagnosis or type of chronic pain, inclusion and exclusion criteria

**Intervention characteristics:**

- Intervention duration, session frequency and length, music type/genre, music selection procedure, method of music exposure, setting, adherence monitoring, control intervention

**Outcomes and results:**

- Outcomes assessed, measurement instruments, and main findings (including data used for the meta-analysis: means, SDs, and sample sizes for pain intensity)

### Data extraction Table

| Study | Country / Design | Sample (n) | Age (years) | Sex | Diagnosis | Intervention (duration; frequency; length) | Music type / selection | Setting | Adherence monitoring | Control | Outcomes | Main results |
|---|---|---|---|---|---|---|---|---|---|---|---|---|
| McCaffrey & Freeman, 2003 | United States; RCT 2-arm | Total 66; M 33; C 33 | >65; range NR | F 44; M 22 | Osteoarthritis | 2 weeks; daily; 20 min | Relaxation classical (Mozart); researcher-selected | NR (likely home) | No | Quiet rest / reading | Pain | Significant reduction in pain and anxiety vs control. |
| Siedliecki & Good, 2006 | United States; RCT 3-arm | Total 60; PM 18; SM 22; C 20 | Mean 49.7; 26–64 | F 46; M 14 | Chronic non-malignant pain | 1 week; daily; 60 min | Standard vs patterned relaxing music; PM self-selected | NR | No | Standard care | Power; Pain; Depression; Disability | Reduced pain, depression, disability vs control; no difference between music types. |
| Guétin et al., 2012 | France; RCT 2-arm | Total 87; M 44; C 43 | Mean 47.8; 19–84 | F 68; M 19 | Musculoskeletal / neuropathic pain | 60 days; 2/day; 20 min | Relaxation music, "U" technique; researcher-guided | Rooms, eyes closed, earphones | Yes (recorded duration/frequency) | Standard care | Pain; Anxiety; Depression; Medication use | Significant reduction in pain, anxiety, depression; reduced medication; effect lasted 1 month post. |
| Onieva-Zafra et al., 2013 | Spain; RCT 2-arm | Total 55; M 28; C 27 | Mean 51.6; 37–65 | F 53; M 2 | Fibromyalgia | 4 weeks; daily; 60 min | Classical mixed with salsa; researcher CDs | Home | Diary (CD usage) | Standard care | Pain; Depression | Significant decrease in pain and depression; reduced stress, increased relaxation. |
| Innes et al., 2018 | United States; RCT 2-arm | Total 22; M 11; C 11 | Mean 58.46; 50–74 | F 15; M 7 | Knee osteoarthritis | 8 weeks; 2/day; 15–20 min | Relaxing instrumental (6 composers); participant choice | Home, quiet, eyes closed | Yes (follow-up calls) | Mantra meditation | Pain; Function; Stress; Mood; QoL | Improved knee function and reduced pain in both groups; reduced stress. |

| Study | Country / Design | Sample (n) | Age (years) | Sex | Diagnosis | Intervention (duration; frequency; length) | Music type / selection | Setting | Adherence monitoring | Control | Outcomes | Main results |
|---|---|---|---|---|---|---|---|---|---|---|---|---|
| Torres et al., 2018 | Spain; RCT 2-arm | Total 56; M 33; C 23 | Mean 50.97; 35–62 | F 56 | Fibromyalgia (women) | 3 months; 1/week; 6–18 min music within 2-h session | Classical; group music & imagery (GrMI) | Soundproof clinical room | No | Standard care / guided imagery | Pain; Well-being; Function; Anxiety; Depression | Reduced pain intensity, improved well-being and state anxiety; sustained at follow-up. |
| Lin et al., 2020 | China; RCT 2-arm | Total 86; M 43; C 43 | Mean 52.3; range NR | F 44; M 42 | Chronic post-surgical (valve replacement) | 6 months; daily; 30 min | Light music, folk, opera, pop; participant choice | Quiet room, 8–10 pm | No | Quiet rest | Pain; QoL | Significant reduction in chronic pain and improved QoL. |
| Eshuis et al., 2021 | Denmark / Italy / Netherlands; RCT 2-arm | Total 45; LF 23; HF 22 | Mean NR; 66–79 | F 33; M 12 | Chronic musculoskeletal pain (elderly) | 3 weeks; 2/day; ~24 min (8 sessions) | Rhythmically aligned music + low-frequency vibration (20–100 Hz) | Standardized clinical room | No | Higher-frequency vibration (200–300 Hz) | Pain; QoL; Pain-related disability | Reduced pain and disability in both groups; no significant difference between frequencies. |
| Du et al., 2022 | China; RCT 2-arm | Total 37; M 19; C 18 | Mean 51; range NR | F 22; M 15 | Chronic (non-oncological) pain | 7 days; daily; 30 min | Self-selected relaxing music (8–150 Hz, 50–70 dB) | NR | Yes (app / software) | Usual care (exercise) | Pain; HRV; Brain hemodynamics | Significant reduction in pain; improved HRV and prefrontal activity. |
| Raglio et al., 2023 | Italy; RCT 3-arm | Total 24; PG 8; MG 8; C 8 | Mean 52.5 / 54; range NR | NR | Fibromyalgia | 1 month; 2/day; 30 min | Preferred playlist vs Melomics-Health algorithmic music | Home | Yes (app) | Standard care | Pain; QoL | Improved well-being in music groups; no significant change in pain perception. |

*M = music group; C = control; PM = patterned music; SM = standard music; PG = preferred (playlist) group; MG = Melomics group; LF = low frequency; HF = high frequency; F = female; NR = not reported; QoL = quality of life; HRV = heart rate variability.*
